# Hyperinsulinemia counteracts inflammation by suppressing IFNγ and inducing senescence in CD4^+^ T cells of patients with rheumatoid arthritis

**DOI:** 10.1101/2024.03.08.24303970

**Authors:** Malin C Erlandsson, Lauri Weman, Eric Malmhäll-Bah, Venkataragavan Chandrasekaran, Mahomud Tuameh, Karin ME Andersson, Sofia Töyrä Silfverswärd, Lisa M Nilsson, Tatiana Zverkova Sandström, Rille Pullerits, Mats Dehlin, Tuulikki Sokka-Isler, Maria I Bokarewa

## Abstract

**Background:** Clinical evidence connects hyperinsulinemia with obesity, and development of type 2 diabetes (T2D). However, its role in autoimmune conditions was questioned. We investigated consequences of hyperinsulinemia for development of T2D and CD4 T cell function in rheumatoid arthritis (RA).

**Methods:** Incident T2D was prospectively studied in two independent RA cohorts and in gout patients matched to RA by age and gender, for 10 years. Effect of hyperinsulinemia and JAK-STAT signaling inhibition (JAKi) in CD4 T cells was studied by integrating transcriptional sequencing with direct effect of insulin, and JAKi on cell proliferation, DNA enrichment, and cytokine production.

**Results:** T2D was 3.2-2.5 times less prevalent in RA compared to gout, particularly in females. Hyperinsulinemia predicted the development of T2D, regardless of metabolic parameters and insulin resistance. Additionally, hyperinsulinemia correlated with the senescence-associated high serum levels of IL6, IL8, and VEGF.

Hyperinsulinemia, along with ex-vivo exposure of CD4 cells to insulin, inhibited cell cycle progression and induced DNA enrichment through the suppression of the PI3K-Src kinases and cell cycle promoting genes. It also reduced IFNγ production. JAKi-treated CD4^+^ cells regained insulin sensitivity, which activated glucose metabolism and facilitated senescence. This insulin-dependent mechanism promoted the accumulation of naïve CD4 cells in JAKi-treated patients.

**Conclusions:** This study shows that insulin has important immunosuppressive ability controlling the adaptive immunity by suppressing IFNγ production and inducing senescence in the effector CD4 T cells. Inhibition of JAK-STAT signaling enhances insulin sensitivity and rejuvenates CD4 cell population in RA patients.

## Introduction

Hyperinsulinemia is known as an early predictor of metabolic dysfunction. Numerous clinical studies connected hyperinsulinemia with obesity, poor lipid profiles and showed its essential role in triggering adverse glucose metabolism and contributing to the development of type 2 diabetes (T2D) [1, 2]. Susceptibility to T2D has gender differences. Men are prevalent among the T2D patients and develop the disease earlier in life compared to women [3]. However, the life-threatening complications of T2D are more prevalent in women.

Epidemiological studies in rheumatoid arthritis (RA) demonstrated contradictive results regarding frequency of T2D development. Solomon et al. reported the increased risk of T2D in RA compared to age- and sex-matched controls [4], while the risk appeared similar between RA and controls in other studies, when adjustment for life style, and metabolic parameters was applied [5, 6]. The later findings are surprising because chronic inflammation in RA is associated with significant production of insulin. Indeed, the prevalence of hyperinsulinemia in RA is significantly higher compared to the general population and to patients with SLE [6–8]. Relationship between inflammation and T2D is clearly bidirectional. On one hand, inflammation induces excessive insulin production, which is frequently seen in severe infection and in critically ill patients [9, 10]. On the other hand, experimental insulin infusion causes activation of leukocytes in serum and adipose tissue followed by IL6, TNFα and MCP1 production in healthy subjects [11, 12], which leaves the physiological role of hyperinsulinemia unresolved.

Sensitivity of leukocytes to insulin is an important determinant in advancing energy supply through glycolytic flux intensity. On surface of immune cells, insulin activates insulin receptor (InsR)/IGF1 receptor (IGF1R) dimer and promotes glucose uptake and cellular growth and differentiation regulating critical aspects of immune cell behavior. Chronically elevated insulin levels decreased insulin signaling, known as insulin resistance, manifesting with a decrease of growth and maturation supporting an anti-inflammatory phenotype of macrophages [13]. In adaptive immunity, insulin signaling is a requisite for a sufficient effector function of T cells [14, 15], the major cellular contributor to RA pathology. Analogously, experimental induction of insulin resistance by either a deletion of InsR [14, 15] or an inhibition of the adaptor IRS proteins [16] reduced the antigen-specific T cell responses including proliferation and cytokine production and deprived T cells of cytotoxic ability. However, knowledge about the direct effect of insulin on CD4^+^ cell function remains limited.

The aim of this study was to prospectively investigate development of T2D in RA patients and to study functional consequences of hyperinsulinemia for CD4^+^ T cells. Development of T2D in RA was compared to gout. Gout was chosen as a positive control group because of the known causality related risk of T2D in these patients [17, 18]. We experimentally verified the effect of insulin on cell cycle progress, senescence, and cytokine production by CD4^+^ cells. We investigated transcriptional differences of insulin signaling in female and male CD4^+^ cells and studied the footprint of chronic hyperinsulinemia on CD4^+^ cells. Finally, we examined the contribution of JAK/STAT inhibition to insulin signaling under experimental conditions and in JAKi-treated RA patients.

### Patients and methods

*RA cohort of Central Finland (RA-FIN)* (Supplementary Figure S1A, S1B). The cohort of patients diagnosed with RA at the Rheumatology unit, Jyväskylä Central Hospital, which conducts the referral specialized ward in the Central Finland with the total population of 250.000 people. Since January 1, 2007, patients with the diagnosis of RA were enrolled in the monitoring system GoTreatIT, which is shared between the specialized and primary health care, and followed according to the clinical protocol.

For purpose of the study, comorbidity data were extracted from the Finnish Rheumatology Quality Register. Among 2987 RA patients, 255 had known DM at RA diagnosis and those 46 with no records after the diagnosis were excluded from the follow-up. A search for incident DM was applied in the total of 2714 patients (1892 female and 822 male) who had completed the period of 10 calendar years after the RA diagnosis by July15, 2023 and were residents of the Central Finland County. New DM case was identified when the diagnostic code (ICD-10 code E10 and E11) and a new drug prescription was registered. Extraction of information from the registry for the purpose of research has been approved by the Wellbeing Services County of Central Finland December 2013 and the amendment in June 2023.

*RA cohort of Göteborg (RA-SW)* (Supplementary Figure S1A, S1B). In total, 328 RA patients (256 female and 74 male) were included in the study at the Rheumatology Clinic of Sahlgrenska University Hospital, Gothenburg, and the Northern Älvsborg County Hospital, Uddevalla during the period between November 2, 2011 and December 14, 2018 [16, 19]. The study is approved by the Swedish Ethical Review Authority and was registered at the Clinical Trials.gov with ID NCT03449589.

In the present study, 19 patients with DM at baseline were excluded, which resulted in the follow-up cohort of 309 patients. At inclusion, the body mass index (BMI) was calculated based on height and weight and expressed in kg/m^2^. Disease activity score (DAS28) of RA was calculated based on assessment of 28 tender and swollen joints and erythrocyte sedimentation rates (ESR). All RA patients fulfilled the EULAR/ACR classification criteria [20] and gave their written informed consent prior to the blood sampling.

The prospective follow-up in RA-SW was conducted during years 2018, 2021 and 2022. The patients were contacted for structured telephone interviews, which included questions regarding DM diagnosis and treatment. The patient report, medical records and the digital prescription lists were used to ratify DM diagnosis. The new cases of DM were further confirmed in the Swedish National Patient Register. In total 6 patients dropped out at follow-up.

Transcriptomics of CD4^+^ T cells was performed on 56 female non-diabetic RA patients. Among these patients, 24 were treated with JAK inhibitors (JAKi) and 32 had no JAKi (Supplementary Figure S1C). Twelve patients combined JAKi with addition of methotrexate (MTX), 4 with other conventional DMARD, 3 with biological DMARD (1 abatacept, 1 tocilizumab, 1 sarilumab). The patients with no JAKi were treated with MTX monotherapy (n=14), biological DMARD (10 TNFα inhibitors, 1 tocilizumab), no active DMARD (n=7). Oral corticosteroids were used by 2 patients in the JAKi-treated group and by 3 in the no-JAKi group.

*Gout patients.* We used the West Sweden Gout registry VEGA [21] to match RA-SWE patients. To be included in VEGA, the patient should have age ≥18 years and diagnosis of gout (ICD code M10) given by a physician. Between the period 2001 and 2010, VEGA comprised 23800 gout patients who were randomly matched to the RA-SWE patients by sex and year of birth at ratio 3-to-1. Patients were excluded if they had a prevalent diagnosis of DM (ICD-10 code E10 and E11) or if they had died or migrated before January 1, 2011. This resulted in 1011 gout patients (female 798, male 213), who were included in the follow-up (Supplementary Fig S1A,S1B). Gout patients were then followed between January 1, 2011 and December 31, 2019. The data collection of gout patients and their comorbidities was approved by the Ethical Review Board of Gothenburg, Sweden.

*Blood sampling and storage.* Blood was collected between 7am and 10am after overnight fasting. For serum and plasma preparation, the blood was obtained from the cubital vein into vacuum containers (BD Vacutainer). Samples were aliquoted and stored at -70°C.

*Serological measurements.* Serum IGF1, total cholesterol (TC), triglycerides, high-density lipoprotein (HDL) and low-density lipoprotein (LDL) were measured by photometry on Cobas 8000 (Roche Diagnostics, Switzerland) at the Laboratory of Clinical Chemistry, Sahlgrenska University Hospital, within routine care of RA patients. Plasma glucose levels were measured using FreeStyle Lite (Abbott Diabetes Care Ltd., Oxon, UK). Insulin levels in plasma was measured by a sandwich ELISAs (DY8056, R&D Systems, Minneapolis, MN, USA). According to the manufacturer, plasma mean±SD insulin levels in fasting healthy people were 60 ± 39 pmol/L. Thus, the insulin levels above 157 pmol/L indicated hyperinsulinemia in this study. C-peptide levels in urine were measured by sandwich ELISA (DICP00, R&D Systems). Insulin resistance was estimated by TyG index, which was the natural logarithm (ln) of (glucose x triglyceride levels in plasma/2) [22].

Rheumatoid factor (RF) and anti-citrullinated protein antibodies (ACPA) were measured in serum samples at the accredited Laboratory of Clinical Immunology at the Sahlgrenska University Hospital. ACPA was measured using an automated multiplex method (Bioplex2200, Biorad, Hercules, CA).

### Cell isolation and culturing

Human peripheral blood mononuclear cells (PBMC) were isolated from the peripheral heparinized blood of healthy controls (26 female, 14 male, age 46±13 years) by density gradient centrifugation on Lymphoprep (Axis-Shield PoC As, Norway). CD4^+^ T cells were isolated by positive selection (Invitrogen, 11331D), and cultured at cell density 1.25x10^6^ cells/ml in wells coated in anti-CD3 antibody coated plates (0.5 μg/mL; OKT3, Sigma-Aldrich, St.Louis, Missouri, USA) in the Roswell Park Memorial Institute (RPMI 1640) medium (Gibco, Waltham, Massachusetts, USA) supplemented with 5% fetal bovine serum (Sigma-Aldrich, St.Louis, MO, USA), 4 mM Glutamax (Gibco), 50 mM β_2_-mercaptoethanol (Gibco), and 50 mg/mL gentamycin (Sanofi-Aventis, Paris, France) at standard conditions of temperature, CO_2_ pressure and humidity. The culture media was supplemented with insulin (0 and 10nM, Humalog 100 U/ml, Eli Lilly, Sweden) for 48h at 37°C and/or tofacitinib (0 or 10 µM, Selleck Chemicals, Houston, TX).

For RNA-seq, CD4^+^ cells were isolated by positive selection (Invitrogen, 11331D) from PBMC of 56 female non-diabetic RA patients (Supplementary Figure S1C) and cultured (1.25x10^6^ cells/ml) for 2 h in wells coated with anti-CD3 antibody (0.5 μg/mL; OKT3, Sigma-Aldrich, St.Louis, Missouri, USA).

### DNA content and cell cycle analysis

CD4^+^ T cells were stained with CellTrace violet (Invitrogen, Waltham, Massachusetts, USA) according to the manufacturer’s instruction to follow cell proliferation and stimulated as above for 48h with or without insulin or a non-selective inhibitor of Janus kinases tofacitinib (JAKi). At harvest, supernatants were collected, cells were permeabilized for 1h, 4°C with cytofix/cytoperm (BD Biosciences, Franklin Lakes, New Jersey, USA) and incubated overnight, 4°C, with 20 μg/ml 7-Aminoactinomycin D (7AAD) (Invitrogen) in perm/wash (BD biosciences). Stained cells were washed 3 times with perm/wash and resuspended in 200 μl FACS buffer. Cells were acquired in FACS Verse (BD Biosciences). Analysis of the acquired data was performed using the Tree Star FlowJo software using the built-in cell cycle analysis tool, Watson model with constrains, CV (G2) = CV (G1) [23]. The CellTrace Violet stain diluted, and the mean fluorescence intensity (MFI) reduced with cell proliferation, while 7AAD-MFI increased with accumulation of DNA content in the cells. The acquired CD4^+^ cells were gated on 7AAD^+^ and separated by the forward cell scatter (FCS) into a large-cell size and small-cell size populations and used for cell cycle analysis resulting in G1, S and G2 phases. DNA content was analyzed in 7AAD^hi^ cells (Q2=LSC and Q3=SSC). (Supplementary Figure S2A).

### Cytokine measurement

The measurement of cytokines in serum and supernatants of CD4^+^ cell culture was performed using sandwich ELISAs for VEGF (DY293B), IL-8 (DY208), IL-10 (DY217B), and survivin (DYC647) (R&D Systems, Minneapolis, MN, USA). IL-6 (M1916) IL1β (M1934) and IFNγ (M1933) (Sanquin, Amsterdam, The Netherlands), as previously described [24, 25].

Dot blot cytokine array Proteome Profiler™ Array, Human Cytokine Array Panel A (ARY005, R&D Systems), which allowed investigating production of 36 cytokines and chemokines in the supernatants of cultured cells was used as described [26]. In short, the array membranes precoated with capture antibodies were soaked in the supernatant of the insulin-stimulated and control cells pooled in equal volume from each cell culture.

### Gene expression analysis by conventional RT-PCR

RNA from CD4^+^ cell cultures was prepared using the micro mRNA kit (Norgen, Ontario, Canada). Complementary DNA was prepared using High-Capacity cDNA Reverse Transcription Kit (Applied Biosystems, Foster city, CA). Quantitative PCR was performed with SYBR Green qPCR Mastermix (Qiagen) using a ViiA™ 7 Real-Time PCR System (Applied Biosystems). Expression was normalized to the reference gene *ACTB*. (TATAA Biocenter, Sweden). Melting curves for each PCR were performed between 60°C and 95°C to ensure specificity of the amplified product. The results are expressed in relative quantity (RQ) to the expression level in the control cells using the ddCt-method.

### Primer Design

Primers were designed in-house using the Primer3 web client (https://primer3.ut.ee/). When applicable, primers were separated by an exon-exon boundary. Amplicon and primer size was limited to 60-150 and 18-24 base pairs, respectively. Melting temperature was set between 60-63°C, max poly-X to 3 and GC-content was limited to 40-60%. Primers suggested by the software was checked in Net Primer web client (https://www.premierbiosoft.com/netprimer/) for possible hairpin and primers-dimer structures. Lastly, correct binding of primers was validated in UCSC In-Silico PCR web client (http://genome.ucsc.edu/cgi-bin/hgPcr) against the GRCh38/hg38 human genome assembly. Primer sequences can be found in Supplementary Figure S2B.

### Transcriptional sequencing (RNA-seq)

RNA from CD4^+^ cell cultures was prepared using the micro mRNA kit (Norgen, Ontario, Canada). Quality control was done by Bioanalyzer RNA6000 Pico on Agilent 2100 (Agilent, St.Clara, CA, USA). Deep sequencing was done by RNA-seq (Hiseq2000, Illumina) at the LifeScience Laboratory, Huddinge, Sweden. Raw sequence data were obtained in Bcl-files and converted into fastq text format using the bcl2fastq program from Illumina. Fastq-files and the processed reads are deposited in Gene Expression Omnibus at the National Centre for Biotechnology Information with the accession code GSE201669.

Fastq RNA-seq files of CD4^+^ cells (GSE138747) of 77 RA patients (54 women and 23 men) [27] were extracted from the Gene Expression Omnibus database. Clinical variables of the patients were kindly provided by Drs. Aridaman Pandit and Weiyang Tao, University Medical Centre Utrecht, the Netherlands (Supplementary Figure S1C).

### Transcriptional sequencing analysis

Mapping of transcripts was done using Genome UCSC annotation for hg38 human genome assembly. The differentially expressed genes (DEGs) were identified by R-studio using the Bioconductor package, “DESeq2” version 1.26.0 [28].

### Bioinformatic analysis

Functional enrichment analysis for the Gene Ontology defined biological processes (GO:BP) and molecular function (GO:MF) was done for the protein-coding DEG (expression >10, nominal p-value <0.05) in Gene set enrichment analysis (GSEA, Broad Institute, USA) and String database (Switzerland). Unsupervised clustering was applied to the DEG followed by a GO:BP and GO:MF searching. GO:BP were limited to those containing <1500 genes from the analyzed category and utilized the false discovery rate (FDR) 5% p-value adjustment. The GO:BP enriched in female, hyperinsulinemia and JAKi-treated CD4^+^ cells were uploaded into Revigo (http://revigo.irb.hr/) to create a semantic plot of common biological processes.

Glycolytic index was calculated using normalized counts (DESeq2) of gene transcripts representing the glycolytic pathway [29]. The index presents average of the standardized gene expression.

### Statistics

The Kaplan-Meier curves and the log-rank Mantel-Cox analysis were used to calculate the instantaneous risk of T2D over the follow-up period as hazard ratios (HR). The receiver operating characteristics (ROC) curves calculated specificity and sensitivity of individual parameters for T2D presented as area under the curve (AUC). Paired comparison between the groups was done by Wilcoxon test and unpaired comparison by the Mann-Whitney U test. Overall risk for hyperinsulinemia was calculated as Odds Ratio (OR) using www.openepi.com. The Spearman’s correlation was performed in Graphpad Prism. Only a few missing values were present in the crossectional RA-SW cohort (highest 5% for DAS28) and those were ignored in the analyses.

For the differential expression analysis, the samples were split at plasma insulin level >157 pmol/L and the groups were compared using DESeq2. The p-values below 0.05 were considered significant. To prepare the Log2 Fold Change heatmap with nominal p-values, the selected genes were collected from the different DESeq2 datasets into new data frames. The heatmaps were prepared using the ComplexHeatmap package [31]. Correlation heatmaps were prepared using the R package Corrplot [32].

## Results

### Development of T2D in RA and gout

To investigate the frequency of incident T2D in RA patients, we performed a prospective observational study in two independent RA cohorts. To place the frequency of T2D in the metabolic context, we have chosen to compare the RA cohorts with gout DM-free patients (Supplementary Figure S1A, S1B). Within the period of 10 years, new T2D was registered in 2.8% (76/2714) of RA-FIN, 3.6% (11/309) of RA-SW patients and in 9.0% (91/1011) of the gout patients, which demonstrated a significantly lower risk of T2D among both RA cohorts compared to gout (RA-FIN, p=e-7, RR=0.31 [0.23-0.42] and RA-SW, p=8.7e-4, RR=0.39 [0.21-0.73]). The log-rank analysis revealed that the T2D-free survival was longer in female with RA compared to gout (Figure 1A). In RA-FIN, the T2D risk was significantly lower in female and male patients compared to gout. In RA-SW, the T2D risk was significantly lower only in female patients (Figure 1A).

**Figure 1.**
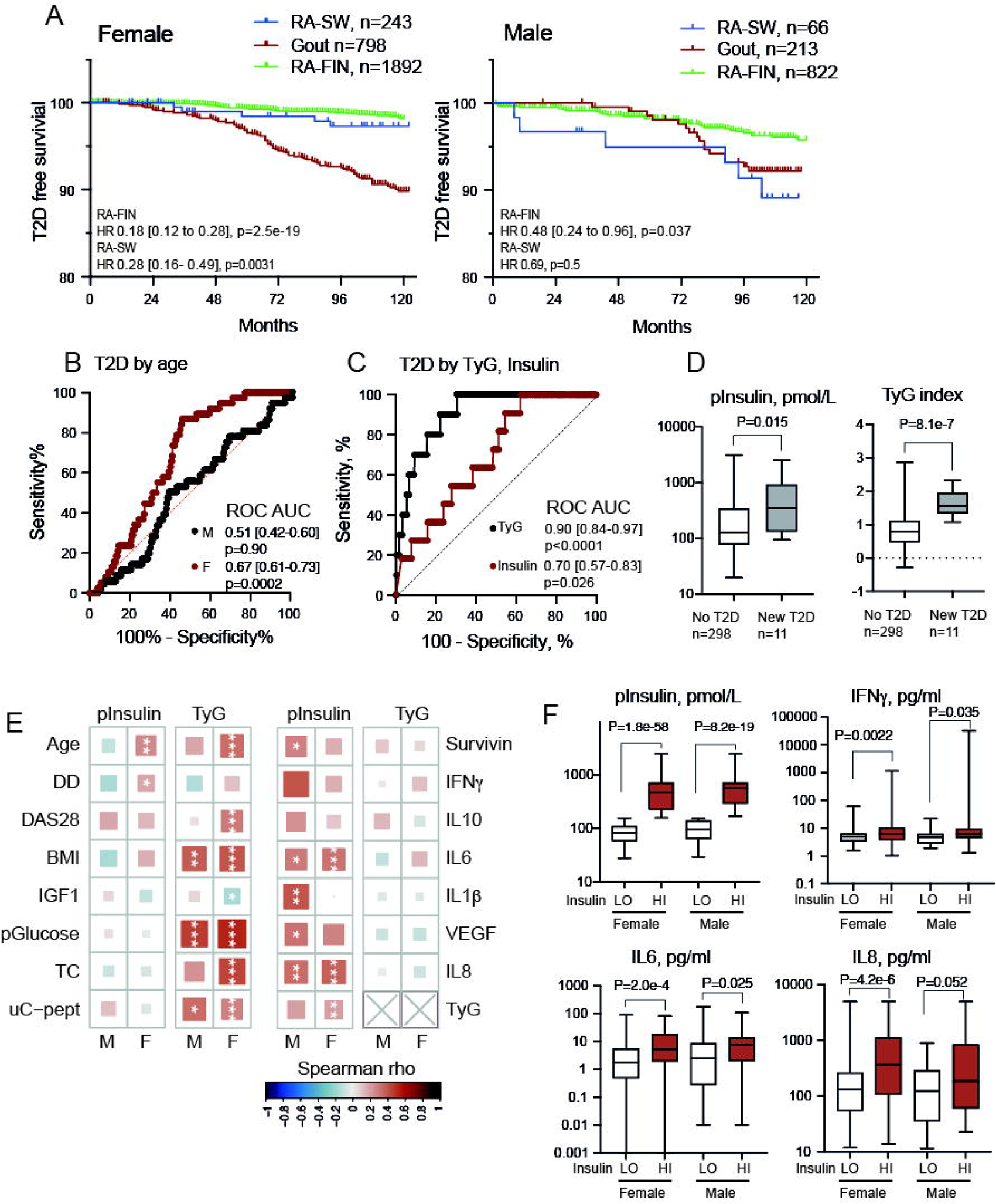
Hyperinsulinemia precedes incidental diabetes mellitus in rheumatoid arthritis. A. Kaplan-Mayer curve of DM-free survival in female and male patients in the Finnish (RA-FIN), and Swedish (RA-SW) and gout cohorts. Risk difference between RA and gout was calculated by the log-rank Mantel Cox test. B. ROC curve analysis of T2D and age at RA diagnosis in RA-FIN and RA-SW patients. Area under the curve (AUC) was calculated by the Clopper method. C. ROC curve analysis of T2D and triglyceride-glucose (TyG) index, and plasma (p)insulin in RA-SW patients. D. Box plots of the baseline pInsulin levels and TyG index in RA-SW patients. P-values are obtained by Mann-Whitney test. E. Heatmap of correlation between RA disease, metabolic variables, and serum cytokine levels with pInsulin and TyG index, in RA-SW patients. * p<0.05, ** p<0.01, ***p<0.001. F. Box plots of plasma (p)Insulin, serum interferon-gamma (IFNγ), IL6 and IL8 levels in RA-SW patients with high (above 157 pmol/L, F, n=81; M, n=39) and low (F, n=122; M, n=27) pInsulin. P-values are obtained by Mann-Whitney test. T2D, type 2 diabetes; DD, disease duration; HR, hazard ratio; IFNγ, interferon-gamma; IGF1, insulin-like growth factor; VEGF, vascular endothelium growth factor; DAS28, disease activity score by 28 joints; BMI, body mass index; TC, total cholesterol; LO, low; HI, high.

To estimate predictive value for incident T2D, we performed the receiver operating characteristic (ROC) analysis of T2D development with respect to age at RA diagnosis, baseline levels of insulin and insulin resistance, by TyG index. The ROC analysis of the regression model showed that age at RA diagnosis was predictive for development of T2D in female patients (Figure 1B). Consistent with previous reports [22, 33, 34], both insulin and TyG index were associated with development of T2D, however the sensitivity of insulin was lower (Figure 1C). Additionally, we found that insulin (p=0.015) and TyG index (p=4.8e-7) were significantly higher in the patients, who later developed T2D (Figure 1D).

Investigating relationship between insulin, TyG index and the RA disease, we found that a relative risk of hyperinsulinemia was significantly higher in females above 60y, DD above 10y, and treatment with methotrexate, which reproduced the high cardio-vascular risk profile specific for RA patients [35, 36]. Unexpectedly, we found a poor correlation between the insulin and TyG index levels, and a strikingly distinct correlation pattern of these variables to metabolic profile (Figure 1E). The TyG index had a strong correlation with the fasting plasma glucose and insulin clearance estimated by the urine C-peptide levels, BMI, and total plasma cholesterol (Figure 1E). These metabolic parameters displayed neither a correlation to insulin nor a difference in patients with hyperinsulinemia (Supplementary Figure S1D). In turn, insulin correlated to pro-inflammatory cytokines IL6, IL8 and VEGF in serum (Figure 1E), and the patients with hyperinsulinemia had significantly higher serum levels of IFNγ, IL6 and IL8 (Figure 1F). Taken together, these results implied a non-traditional biological role of insulin in the context of inflammation. Considering the opposing associations between insulin, inflammation, and the low risk of T2D in RA female patients, one could anticipate gender dependent differences of insulin effects on leukocytes.

### Distinct insulin sensitivity of female and male CD4^+^ cells

To investigate further the gender-dependent differences in the insulin signaling pathway in CD4^+^ cells of RA patients (54 female and 23 male), we used the transcriptional sequencing.

The cells originated from the female and male patients comparable in age, BMI, DAS28, WBC and platelet count (Supplementary Figure S1C).

Despite the similarity in demographics and RA disease activity, the female CD4^+^ cells had downregulation of the InsR pathway (Figure 2A) including *INSR* and the down-stream mediators *IRS1, IRS2,* and *AKT1,* while the levels of *IGF1R* mRNA were similar (Figure 2B). Consequently, the female CD4^+^ cells had low *SLC2A1* transcripts to the glucose transporter GLUT1 and the glycolytic enzymes *G6PD, HK3, PFKFB3, PFKFB2, ALDOA, PGM1, LDHA, PGAM1, ENO1* and *GAPDH,* summarized in the glycolytic index of CD4^+^ cells (Figure 2C). These results suggested that the female CD4^+^ cells had the transcriptional pattern of an intrinsic insulin resistance, while the male CD4^+^ cells appeared to be more sensitive to insulin.

**Figure 2.**
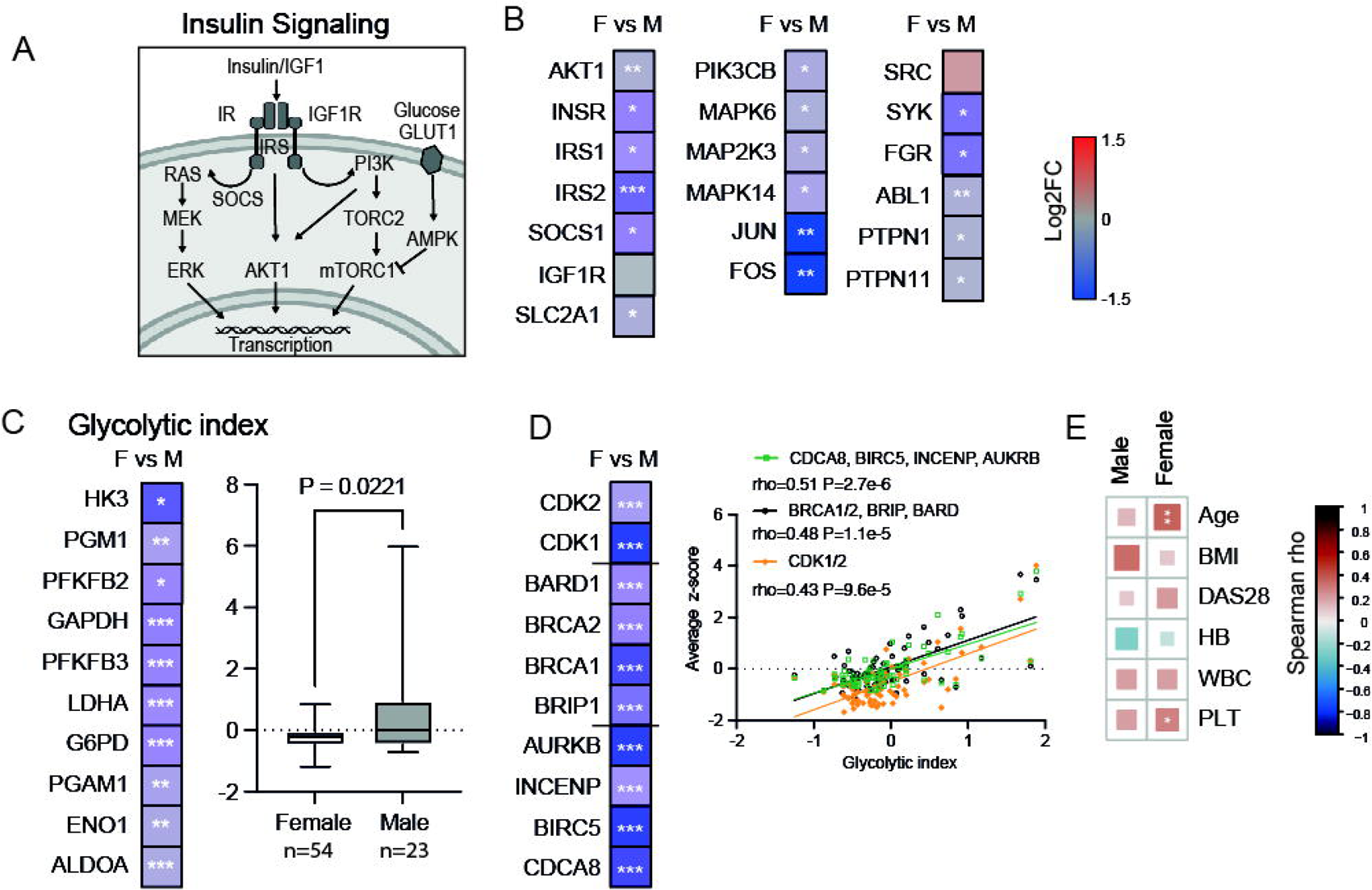
Hyperinsulinemia counteracts inflammation in rheumatoid arthritis. A. Insulin signaling pathway. B. Heatmap of the log2fold change difference in gene expression of female (F, n=54) and male (M, n=23) CD4^+^ cells, by RNA-Seq. P-values are obtained by DESeq2 test. C. Heatmap of expression difference for individual genes and box plot of total glycolytic cell index of CD4^+^ cells, by RNA-Seq. P-values are obtained by DESeq2 test. D. Heatmap of expression difference for genes of the cell division machinery, by RNA-Seq. Dot plot of Spearman’s correlation between glycolytic index and transcription of cell cycle controlling genes. E. Heatmap of the Spearman correlation rho-values between glycolytic index of CD4^+^ cells and clinical parameters. * p<0.05, ** p<0.01, ***p<0.001. BMI, body mass index; DAS28, disease activity by 28 joints; HB, hemoglobin; WBC, white blood cell count; PLT, platelets.

The suppressed insulin signaling was coupled to downregulation of MAP-kinase p38 coded by *MAPK14*, its activator *MAP2K3*, which in cooperation with the AP-1 transcription complex consisting of *JUN* and *FOS,* monitor the cell cycle and DNA metabolism [37, 38] (Figure 2D). Indeed, the female CD4^+^ cells had significant inhibition of the cell cycle kinases *CDK1* and *CDK2*, proteins of the chromosomal passenger complex *AURKB*, *INCENP, CDCA8* and *BIRC5*, and the BRCA1/BRCA2 complex that sensed the DNA double-strand breaks and counteracts senescence [39] (Figure 2F). The glycolytic index had a strong positive correlation with the cell cycle kinases, DNA damage sensing and the chromosomal passenger complex (Figure 2F), which pointed on the insulin reliance of these processes. In contrast, the glycolytic index of CD4^+^ cells had no or only marginal correlation to BMI, DAS28 and WBC count (Figure 2E).

The PI3-kinase signaling was negatively affected through the repression of *PIK3CB* coding for p110 catalytic unit, and the tyrosine kinases *ABL1, SYK*, *FGR, PTPN1,* and *PTPN11* (Figure 2B), which collectively mediated *Cellular Response to Cytokine*s (GO:0071345, FDR 1.04e-39), *Fc-receptor signaling* (GO:0038093, FDR 6.06e-10) and the *Cell Motility* (GO: 0048870, FDR 2.3e-8).

### Insulin inhibits Th1 phenotype and induces senescence in CD4^+^ cells

Based on the results above, which demonstrated an association between transcription of the insulin signaling and cell cycle genes, and the recently reported senescence-inducing properties of insulin [23], we investigated functional effect of insulin on DNA enrichment and proliferation of anti-CD3 activated CD4^+^ cells of healthy individuals, using flow cytometry.

By gating cells stained with the DNA dye 7AAD, we found that 7AAD^+^CD4^+^ cells consisted of the large-size cell (LSC) and a small-size cell (SSC) subsets (Supplementary Figure S2A). Between these two subsets, the SSC had a significantly lower DNA content, which was also significantly increased in response to insulin (Figures 3A). The DNA content remained largely unchanged in the LSC. Insulin stimulation had no significant effect on the proliferation rate of the cultured CD4^+^ cells visualized by the CellTrace violet content (Supplementary Figure S2C). To estimate when during the cell cycle the insulin-induced increase of DNA content occurred, we performed cell cycle analysis of SSC subset. This analysis revealed that insulin induced a significant accumulation of 7AAD^+^ cells in the G1 phase obstructing their transition to S phase, which was reduced (Figure 3B).

**Figure 3.**
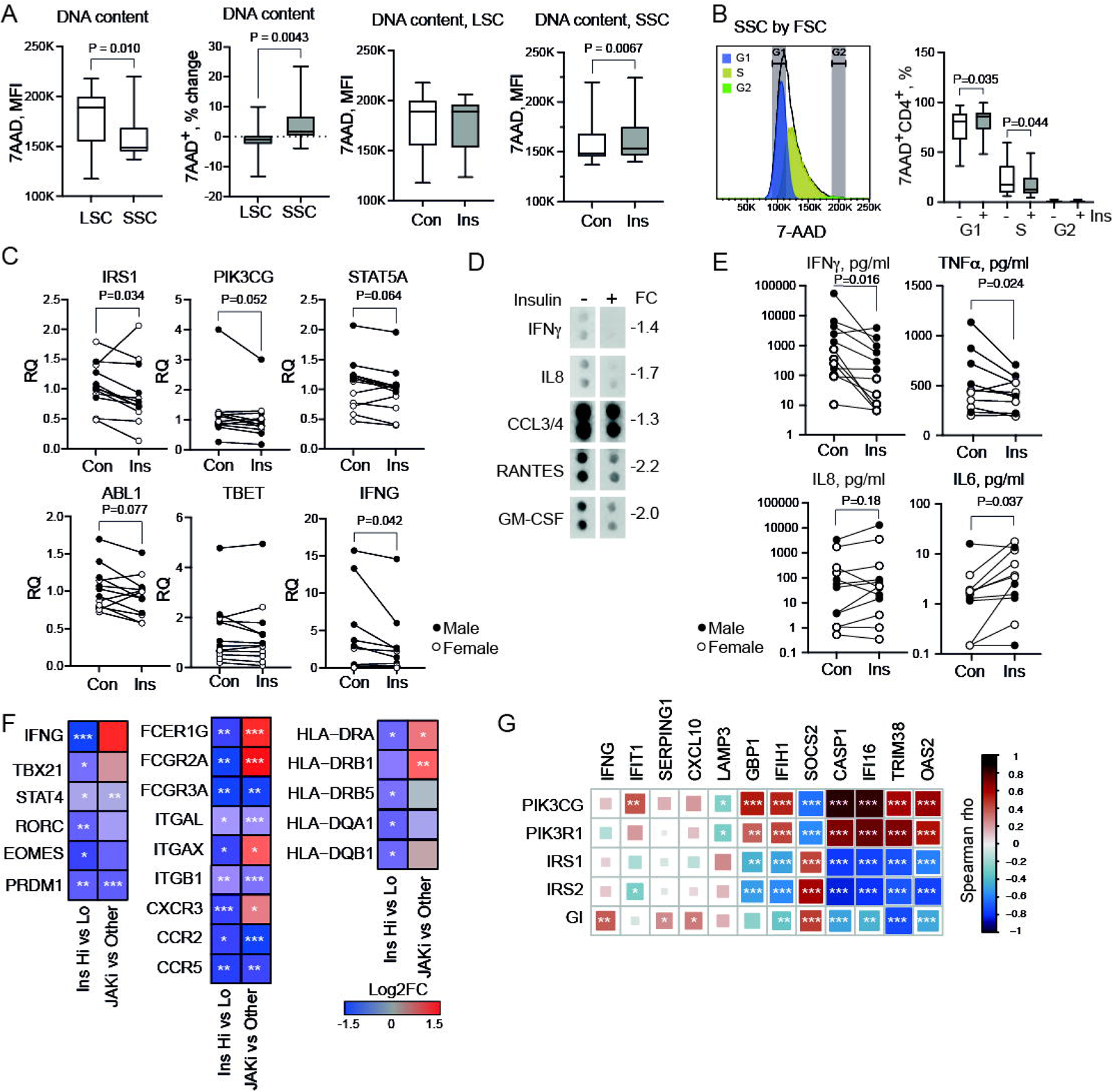
Insulin suppresses IFNγproduction and induces senescence in CD4^+^ cells. CD4^+^ cells (11 females, 7 males) stimulated with anti-CD3 and insulin 10 nM for 48 h were analyzed by flow cytometry. DNA content (7AAD) and cell size (FSC) separated large-size cell (LSC) and small-size cell (SSC) subsets. P-values are obtained by paired Wilcoxon test. A. Box plot of DNA content by mean fluorescence intensity (MFI) of 7AAD^+^ cells. Box plot of DNA content change with insulin treatment. Con, control; Ins, insulin treated cells. B. Histogram of 7AAD^+^ cell distribution by phases of the cell cycle. Colored areas indicate cells in G1 (blue), S (yellow) and G2 (green) phases. Box plot of frequency of 7AAD^+^ cells in different phases of cell cycle. C. Dot plot of gene expression, by qPCR, in relative quantity (RQ) to mock treated cells. D. Dot blot images of cytokine levels in pooled (6 male) supernatants of insulin-treated and control cells. FC, fold change. E. Dot plot of cytokine protein levels in the supernatants, by specific immunosorbent assay. F. Heatmap of gene expression difference by log2 fold change, by RNA-Seq. Samples are grouped by high (n=12) and low (n=44) insulin, and by JAKi-treated (n=24) and others (n=32), P-values are obtained by DESeq2 test. G. Heatmap of Spearman correlation rho-values between expression of IFN-signature genes, insulin signal mediators, and glycolytic index (GI). * p<0.05, ** p<0.01, ***p<0.001.

Investigating direct insulin effect on transcription and protein products of CD4^+^ cells, we found that insulin stimulation repressed *IRS1*, *PIK3CG*, *STAT5A,* and *ABL1* mRNA levels, which was consistent with development of an insulin resistant state in these CD4^+^ cells (Figure 3C). The transcriptional response was detectable in male CD4^+^ cells (Supplementary Figure S2E), while no transcriptional change was found in the female CD4^+^ cells. These transcriptional results of low insulin sensitivity of female CD4^+^ cells supported the observations done in RA patients (Figure 2).

The effect of insulin on cytokine protein production was assessed by screening supernatants of CD4^+^ cells with a cytokine dot array. It demonstrated that the insulin stimulation inhibited production of IFNγ, IL8, CCL3/4, RANTES, and GM-CSF (Figure 3D). Consistent with the results of the dot array and transcriptional suppression of *INFG* gene (Figure 3C), enzyme-linked sandwich ELISA demonstrated that insulin significantly suppressed IFNγ and TNFα production, and increased IL6 in supernatants of CD4^+^ cells (Figure 3E).

Together, these functional results demonstrated the direct effect of insulin on CD4^+^ cells causing DNA enrichment and cell cycle stagnation in the G1 phase, which was also associated with an increase of IL6 production characteristic for senescent phenotype of these cells.

### Chronic hyperinsulinemia is associated with inhibition of the adaptive immune response in CD4^+^ cells

To further investigate the insulin-induced inhibition of IFNγ production presented above, we assessed how chronic hyperinsulinemia affected CD4^+^ cells. We compared the transcriptional profile of CD4^+^ cells of 12 non-diabetic female RA patients with hyperinsulinemia and 44 patients having no hyperinsulinemia. This analysis revealed that hyperinsulinemia was frequently associated with downregulation of gene transcription in CD4^+^ cells (836/1172, 71%. Supplementary Figure S3A). A search for biological processes enriched among the downregulated genes revealed that their role in *Regulation of Immune Processes (*GO:0002682), *Positive Regulation of T cell activation* (GO:0050870) and *Response to* IFNγ *signaling* (GO:0034341) (Supplementary Figure S3B).

The findings in the pathway enrichment analysis were consistent with the insulin-induced suppression of IFNγ in CD4^+^ cell cultures. Indeed, the *IFNG* gene was the top gene repressed in CD4^+^ cells of patients with hyperinsulinemia. Additionally, hyperinsulinemia caused a significant repression of the TFs *STAT4, TBX21*, *RORC, PRDM1*, and *EOMES* as well as receptors assisting induction of IFNγ production including Fc-receptors, integrins, MHC-II receptors and chemokine receptors (Figure 3F). Together, it provided molecular basis for the IFNγ production arrest and argued for a reduced frequency of Th1 cells in patients with hyperinsulinemia.

The inhibition of the Th1 cell profile was followed by the repression of the IFN-stimulated genes clinically important in RA and assembled in IFN signature [40, 41]. Analyzing the IFN-signature genes, we found their uneven expression in hyperinsulinemia and a heterogenic correlation with insulin signaling mediators IRS1/2, glycolytic index, and PI3-kinases (Figure 3G).

An independent cluster of the genes differentially expressed in hyperinsulinemia was annotated to the *Cell Cycle* pathway (GO:0007049, FDR 2.39e-6) (Figure 4A). This included upregulation of cyclin-dependent kinases *CDK2, CDK4*, and their inhibitor *CDKN2D* controlling G1/S transition (Figure 4B), which was experimentally affected by insulin in CD4^+^ cell cultures (Figure 3B), and a repression of *CDK1* essential for G2/M transition of cell cycle. Additionally, hyperinsulinemia inhibited transcription of the cohesion proteins *MELK, CDCA7, ESCO2*, mitotic spindle assembly *KIF15, KIF13A, SGO1,* and *AURKB*; kinetochore proteins *KNL, NDC80* (Figure 4B), which made the cells susceptible to cell cycle arrest and senescence shown in the *in vitro* experiments. Notably, inhibition of the cell cycle genes was the feature common between CD4^+^ cells exposed to hyperinsulinemia and female CD4^+^ cells (Figure 2).

**Figure 4.**
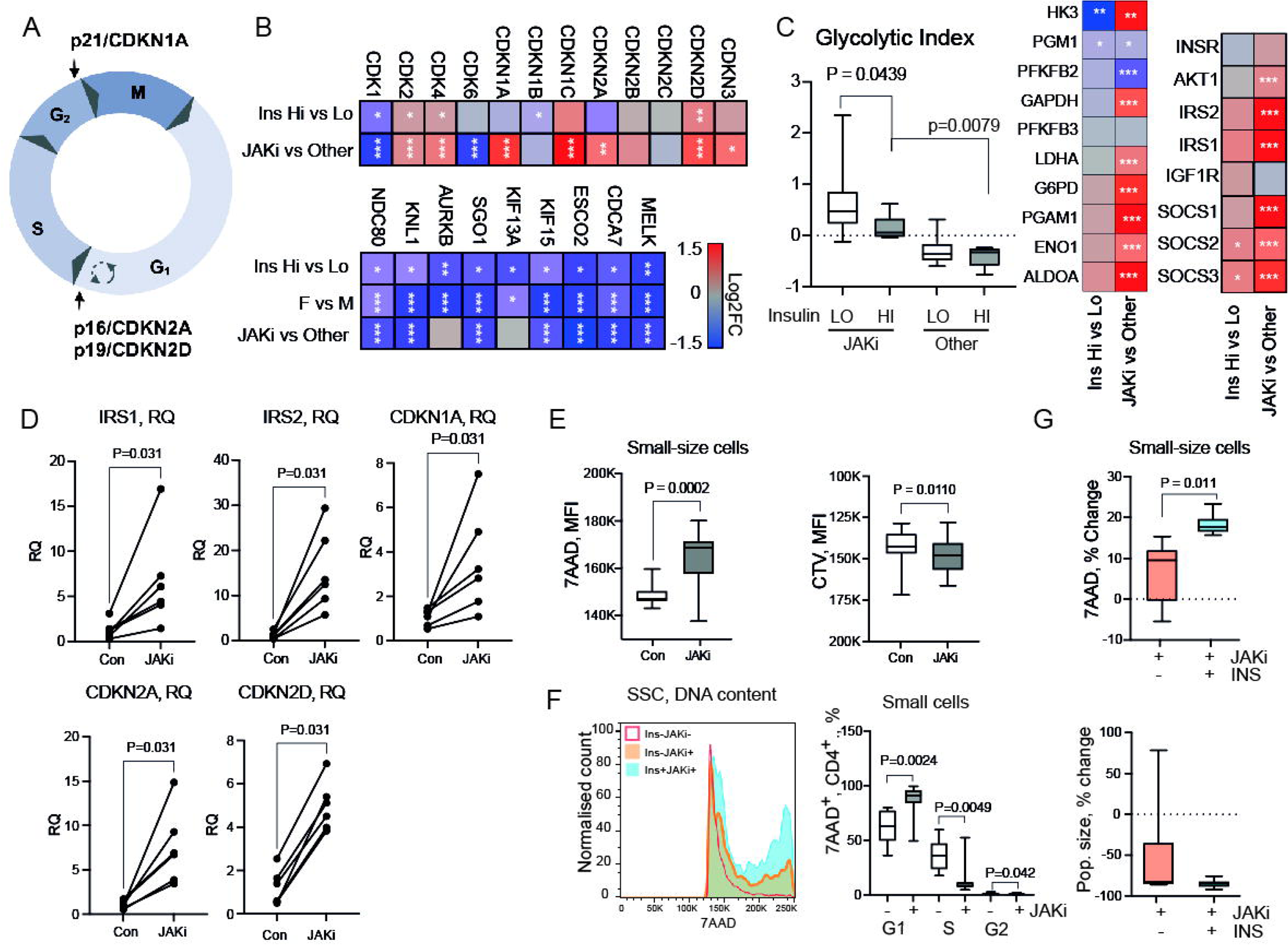
JAK-inhibitors increase insulin sensitivity and aggravate senescence in CD4+ T cells. A. Cell cycle phases and check points control. CD4^+^ cell transcriptome of RA patients with high (n=12) and low (n=44) plasma insulin, JAKi-treated (n=24) and other (n=32), female (n=54) and male (n=23), by RNA-Seq. Difference between groups was analyzed by DESeq2-test. * p<0.05, ** p<0.01, ***p<0.001. B. Heatmap of transcription difference by log2 fold change (log2FC) in cyclin-dependent kinases and their inhibitors. Heatmap of transcription difference in cell division machinery genes. C. Heatmap of transcription difference in genes of insulin signaling and enzymes of the glycolytic index. Box plot of glycolytic index in JAKi-treated (Hi, n=7, Lo, n=17) and others (Hi, n=5, Lo, n=27), split by insulin. D. Dot plot of gene transcription in CD4^+^ cells (n=6) stimulated with anti-CD3 and tofacitinib (JAKi, 10 microM, Con, 0 microM) for 48 h. mRNA levels were analyzed by qPCR and presented in relative quantity (RQ) to mock treated cells. E. Box plot of DNA content by mean fluorescent intensity (MFI) in control and JAKi treated cells. F. Histogram of 7-aminoactinomycin D (7AAD)^+^ cells distribution in different phase of cell cycle, gated on small size cells (SSC) by forward cell scatter. Colored areas indicate cells in G1 (blue), S (yellow) and G2 (green) phases. Box plot of frequency of 7AAD^+^ cells in different phases of the cell cycle. G. Box plot of DNA content change by 7AAD and population size change in cultures stimulated with JAKi and insulin (10 nM, n=6). P-values by paired Wilcoxon test.

### Abrogation of JAK/STAT signal promotes insulin signaling and induces senescence in CD4^+^ T cells

Among the analyzed sets of CD4^+^ cells, 24 belonged to the RA patients treated with JAK-inhibitors (JAKi) that elicited anti-inflammatory effects by abrogating signaling from the common γ-chain cytokine receptors [42]. Thus, we asked if JAKi interfered with the insulin effects on CD4^+^ cells. Comparing CD4^+^ cells of JAKi-treated (n=24) and non-JAKi-treated patients (n=32), we found a significant upregulation of the insulin signaling genes including *IRS1, IRS2, AKT1* (Figure 4C), which denoted improvement of the insulin sensitivity. Consequent with the improved insulin sensitivity, CD4^+^ cells of JAKi-treated patients had high glycolytic index despite the comparable plasma insulin levels in those patients (Figure 4C, Supplementary Figure S2D). JAKi-treated patients had activation of the senescence controlling genes of CDK2, CDK4, and CDK inhibitors *CDKN1A/*p21*, CDKN1C/*kip2*, CDKN2A/*p16, and *CDKN2D/*p19, while the cell cycle promoting CDK1, and cell division machinery of mitotic proteins remained repressed (Figure 4B).

Departing from the findings in CD4^+^ cells of the JAKi-treated patients, we asked if these transcriptional changes were directly resulted from the JAK/STAT signaling inhibition in the cells. To address this, we cultured anti-CD3 activated CD4^+^ cells in presence of JAKi. Analysis of the JAKi exposed CD4^+^ cells with qPCR demonstrated an increase in *IRS1* and *IRS2* mRNA levels that was consistent with the improved insulin sensitivity observed in CD4^+^ cells of the JAKi-treated patients (Figure 4D). Transcripts of *CDKN1A, CDKN2A,* and *CDK2D* were all significantly increased (Figure 4D). Tracking the DNA content of CD4^+^ cells by 7AAD dye and the proliferation by CTV, we revealed a significant accumulation of DNA content in the SSC of the JAKi-treated cultures compared to the control mock-treated cultures (Figure 4E). The cell cycle analysis of SSC cells demonstrated that the accumulation of 7AAD^+^ cells occurred in G1 and G2 phase (Figure 4F). Additionally, JAKi significantly suppressed the CTV dilution which revealed lower proliferation rate in both the LSC and SSC subsets, which corresponded to low expression of cell division machinery genes in JAKi-treated patients (Figure 4B). Together with the increase in the DNA content, suppression of the proliferation, and activation of CDKN expression corresponded to a senescent phenotype of the JAKi-exposed cells. Culturing of CD4^+^ cells with JAKi and insulin, we demonstrated their synergistic effect on the DNA content accumulation in the SSC (Figure 4G). These experimental results fully supported the CD4^+^ cell transcriptomic of JAKi-treated patients and warranted a direct engagement of JAK/STAT signaling in insulin sensitivity. Importantly, the total cell number in JAKi-treated cultures was reduced (Figure 4G) and the anti-proliferative effect of JAKi remained unchanged after addition of insulin, which implied a gradual elimination of the senescent cells.

To investigate if the additive effect of insulin and JAKi was traceable in CD4^+^ cells of JAKi-treated patients, we compared the effect of hyperinsulinemia on CD4^+^ cells within the JAKi-treated (hyperinsulinemia, n=7, others, n=17) and non-JAKi-treated groups (hyperinsulinemia, n=5, others, n=27), we found an obvious suppressive effect of hyperinsulinemia on the glycolytic index in both patient groups (Figure 4C). Accordingly, hyperinsulinemia maintained its immunosuppressive effect in CD4^+^ cell of the JAKi-treated patients by mitigating an adequate upregulation of the cell division machinery genes, the key Th1 TFs *RORA, RORC, PRDM1*, and *STAT4* (Figure 5A). Consistent with the hypothetic elimination of senescent cells, we found no increase of IL6 and IL8 in serum of JAKi-treated patients with hyperinsulinemia, while the IL6 and IL8 increase was significant in non-JAKi-treated patients (Figure 5B). Accordingly, serum survivin, a proxy of cytolysis, was increased in JAKi-treated patients (Figure 5B). However, hyperinsulinemia could neither suppress the rich IFNγ and IL10 production of CD4^+^ cells of JAKi-treated patients (Figure 5C) nor the dominance of the naïve T cell markers *CCR7, IL2RG*, and *CD27*, the TCR complex genes *CD4, CD3E, CD5, CD6, ZAP70* (Figure 5D). In contrast, the memory T cell receptors *CD44, CD69, IL2RA,* and *CD28* remained low. Thus, we concluded that JAKi treatment and insulin had a traceable additive effect inducing senescence of Th1 cells. Additionally, JAKi facilitated rejuvenation of the circulating T cell population.

**Figure 5.**
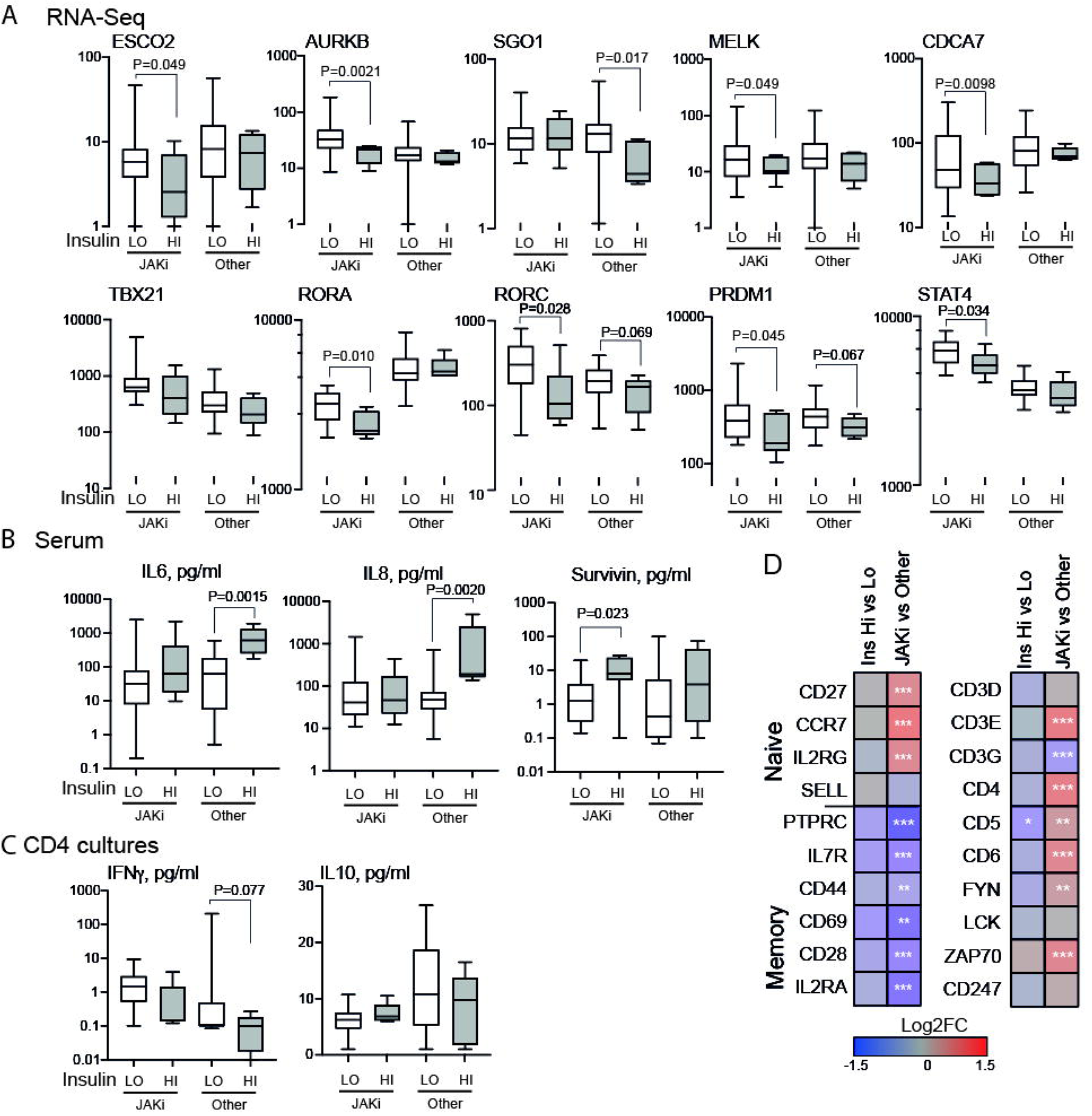
Insulin mitigates transcriptional effect of JAK-inhibitors in CD4^+^ T cells. CD4^+^ cells of female RA patients were isolated by negative selection, activated on anti-CD3 plate for 2h, and analyzed by RNA-Seq. Serum and supernatants of CD4^+^ cell cultures were used for protein measurements by immunosorbent assay. Differences between the groups were analyzed by DESeq2 test and Mann-Whitney statistics. A. Box plot of normalized gene transcription in JAKi-treated (Hi, n=7, Lo, n=17) and other patients (Hi, n=5, Lo, n=27), split by high (Hi) and low (Lo) insulin levels. B. Box plot of IL6, IL8 and survivin levels in serum of patients grouped as above. C. Box plot of protein levels of IFNγ and IL10 in supernatants of CD4^+^ cell cultures. D. Box plots of plasma insulin levels in JAKi-treated (Hi, n=7, Lo, n=17) and other patients (Hi, n=5, Lo, n=27), split by high (Hi) and low (Lo) insulin levels. P-values obtained by Mann-Whitney statistics. E. Heatmap of transcription difference by log2 fold change (log2FC) in gene of the naïve and memory phenotype and TCR specific genes, by RNA-Seq.

## Discussion

In this study, we demonstrated that the risk of T2D in RA was lower compared to gout. The difference was particularly pronounced in female RA patients. Notably, hyperinsulinemia, generally considered as a pre-diabetic state, could explain only a minor part of the T2D risk in RA lacking association between insulin resistance and metabolic parameters. Instead, it exerted strong immunosuppressive properties on effector Th1 CD4^+^ cells and the cell cycle control.

In this study, we demonstrated that IFN-related biological processes were specific targets of insulin. Culturing of CD4^+^ cells and chronic hyperinsulinemia showed a direct suppressive effect of insulin on IFNγ production combined with a significant repression of the key Th1 TFs and the receptors mediating IFNγ production underpinning T cell anergy and senescence observed in *in vitro* experiments. Alternative one-step mechanism of immunosuppression exerted by insulin could engage a direct transcriptional repression of developmental genes. It has recently been shown that, following insulin binding, InsR is transported to the cell nucleus, where it binds RNA polymerase complex and triggers genome-wide changes in gene expression [43]. Notably, insulin resistance abridged accumulation of InsR on chromatin. This transcriptional effect of InsR on the chromatin of T cells awaits further investigation.

Extensive studies of the IFN response in canonic autoimmune diseases of RA, SLE, T1D and multiple sclerosis resulted in a list of IFN-signature genes [44, 45]. In RA, the IFN-signature genes are expected to predict disease progress, development of skeletal damage, and efficacy of antirheumatic drugs [40, 41]. Our study demonstrated heterogenic relation between the IFN-signature genes and hyperinsulinemia, which stressed the role of biological context. Information regarding hyperinsulinemia and insulin sensitivity of CD4^+^ cells appeared to be of relevance for selection of the IFN-signature genes. This knowledge could improve prediction performance of the IFN signature.

The biological forces causing chronic hyperinsulinemia in RA remain unresolved. Results of several studies imply that IFNγ itself is one of the major inducers of hyperinsulinemia. A Finnish pilot study was the first to demonstrate that bolus infusion of IFNα increased plasma insulin levels in healthy subjects, prolonged insulin clearance, and induced insulin resistance [46]. A recent clinical study demonstrated that IFNγ production during viral infection caused insulin resistance in muscles by downregulating InsR, which resulted in an increased insulin secretion by pancreas to balance the insulin resistance [47]. Furthermore, IFNγ deficiency improved insulin sensitivity in mice fed with a diabetogenic diet [48]. Improving insulin sensitivity with biguanides has been shown effective in alleviating experimental arthritis through balancing Th1/Th17 cells [49, 50]. Importantly, the use of biguanides and dipeptidyl peptidase-4 was associated with lower risk for RA development in population-wide studies in USA, United Kingdom, and Taiwan [51–53].

In this study, we showed that abrogation of IFN signaling at the level of JAK-STAT promoted insulin sensitivity of CD4^+^ cells and deepened insulin effects by enhancing cell glycolysis, both *in vivo* and *in vitro*. The regained insulin sensitivity was associated with a general activation of transcription, better cell cycle and DNA damage control. Optimal insulin sensitivity is generally known to counteract insulin production by pancreas [54]. Thus, we hypothesize that elimination of the effector T cells by means of senescence presents a strong protective mechanism that could level-off hyperinsulinemia in RA patients.

Experimental targeting the JAK-STAT pathway achieved a context dependent effect on the energy expenditure, adiposity, insulin sensitivity, and inflammation [55]. Sporadic reports suggest a JAKi-mediated inhibition of glycated hemoglobin level, and improved insulin sensitivity in RA patients with T2D [56] and in non-diabetic patients [57]. This study convincingly demonstrated that JAKi treatment activated insulin signaling in CD4^+^ cells, which prompted T cell anergy and senescence. Furthermore, the JAKi treatment was associated with rejuvenation of the circulating T cell population in RA patients by increasing proportion of the naïve CD4^+^ cells presumably through elimination of senolytic cells. This effect of JAKi partly synergized with hyperinsulinemia, which postponed differentiation of the naïve T cells by suppressing availability of the TFs required for lineage maturation.

We demonstrated that female and male RA patients diverted in T2D development. Potentially, we may see a connection between the gender-dependent difference in insulin sensitivity of CD4^+^ cells and T2D risk difference. The female patients were more resistant to T2D development and had CD4^+^ cells with low expression of the insulin signaling genes and glycolytic index characteristic of insulin resistance. Hypothetically, low insulin sensitivity protects CD4^+^ cells from the immunosuppressive effects of insulin, which retains the effector activity of CD4^+^ cells and booster autoimmunity. In the reverse, efficient autoreactive CD4^+^ cells consume energy, which prevents metabolic alterations in the insulin-sensitive tissues as fat, muscles, liver, and brain in female RA patients. Our observations contribute to the ongoing debate regarding sex related divergency of molecular mechanisms of insulin resistance [58, 59].

Taken together, this study demonstrated that in RA, insulin was disconnected from its traditional metabolic function and had important immunosuppressive ability restricting activity of the effector Th1 cells and facilitating their elimination by means of senescence. Our study has limitations. First, the number of new T2D cases is limited to less than 100 patients despite a sufficient study population and a 10 year-long follow up period, which could comprise a hidden selection bias. Second, the difference in T2D risk observed between female and male RA patients could not be directly translated into age and gender matched healthy controls. This is because of the female dominance in RA disease and the role of IFNγ-related processes attributed to both insulin sensitivity in RA. Third, the study does neither address the effect of oral corticosteroids and other anti-rheumatic treatment on the T2D risk, nor it provides an answer if insulin sensitivity of CD4^+^ cells strengthen the effect of anti-rheumatic drugs in diabetic and non-diabetic RA patients. These questions should be challenged and confirmed in prospective interventional studies and in a variety of autoimmune pathologies.

## Data Availability

This study includes publically available transcriptional data which is deposited at the Gene Expression Omnibus at the National Centre for Biotechnology Information, accession numbers GSE201669 and GSE138747. Other data from the present study are available upon request to the authors

https://www.ncbi.nlm.nih.gov/geo/

## Acknowledgements

We would like to thank the research nurses Anneli Lund and Marie-Louise Andersson at the Rheumatology Clinic, Sahlgrenska University Hospital, Gothenburg, for their help with blood sampling. We also thank all RA patients, who participated in this study. We appreciate support of Aridaman Pandit and Weiyang Tao at the Center for Translational Immunology, University Medical Center Utrecht, The Netherlands, for sharing clinical data of the patients included in GSE138747.

