## Supplementary figure 1 - 3 for "Hyperinsulinemia counteracts inflammation by suppressing IFNγ and inducing senescence in CD4^+^ T cells of patients with rheumatoid arthritis"

A Clinical material

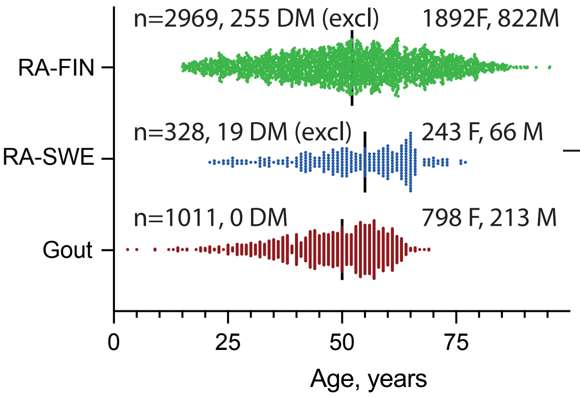


Transcriptional sequencing

RA GSE138747
n=77, 54F, 23M

RA GSE201669
n=60, 4 DM (excluded)
56F

CD4 cultures

Healthy controls
n=40, 0 DM,
26 F, 14 M

B Clinical characteristics of RA and gout patients at baseline

| | RA-FIN Male n=822 | Female n=1892 | RA-SW Male n=66 | Female n=243 | Gout Male n=213 | Female n=798 |
|---|---|---|---|---|---|---|
| Age, y (mean) | 55.0 | 50.2 | 48.0 | 53.1 | 48.2 | 47.5 |
| DD, y (mean) | 0 | 0 | 12.2 | 10.8 | 0 | 0 |
| Follow-up month, n median (range) | 120 (95-120) | 120 (114-120) | 111 (32-117) | 111 (15-122) | 117 (19-117) | 117 (5-122) |

C Clinical characteristics of RA patients in transcriptional sequencing (mean ± SD)

| GSE138747 | Male n=23 | Female n=54 |
|---|---|---|
| Age, y | 50.4 ± 12.2 | 53.7 ± 11.4 |
| DD, y | 0 | 0 |
| BMI, kg/m$^2$ | 26.8 ± 4.1 | 26.7 ± 5.2 |
| DAS28 | 4.11 ± 1.24 | 4.62 ± 1.23 |
| DAS28>3.2 | 18 (78%) | 44 (81%) |
| Hb, mg/L | 144 ± 10.3 | 133 ± 10.7 |
| WBC, 10$^9$/L | 8.07 ± 3.21 | 7.90 ± 3.14 |
| Platelets, 10$^9$/L | 288 ± 57 | 287 ± 58 |

| GSE201669 | JAKi n=24 | Other n=32 |
|---|---|---|
| Age, y | 52.0 ± 12.9 | 64.8 ± 7.7 p<0.0001 |
| DD, y | 15.3 ± 10.4 | 12.2 ± 10.0 |
| BMI, kg/m$^2$ | 26.0 ± 3.6 | 26.2 ± 4.5 |
| DAS28 | 2.35 ± 0.96 | 2.78 ± 1.09 |
| DAS28>3.2 | 3 (9%) | 9 (28%) |
| Hb, mg/L | 132 ± 6.7 | 139 ± 10.6 p=0.033 |
| WBC, 10$^9$/L | 5.4 ± 1.7 | 6.1 ± 2.4 |
| Platelets, 10$^9$/L | 308 ± 88 | 253 ± 57 p=0.025 |

D

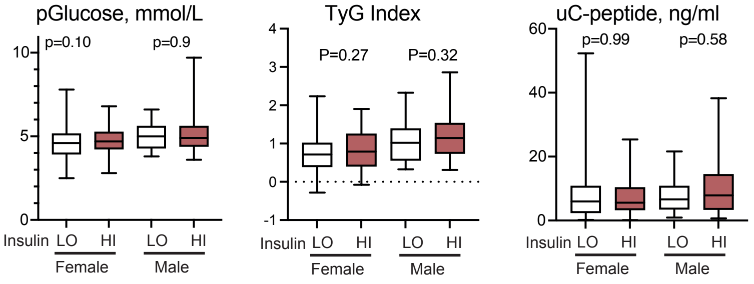


**Supplementary Figure S1.**

A. Histogram of age distribution in the Finnish RA cohort (RA-FIN), Swedish RA cohort (RA-SWE, and gout cohort. DM, diabetes mellitus. F, female, M, male. GSE, Genomic Spatial Event.

B. Clinical characteristics of the patient cohorts.

C. Clinical characteristics of RA patients used in transcriptome analysis of CD4+ cells by RNA-seq. Values are presented as mean+SD. P-values are obtained by Mann-Whitney statistics. DD, disease duration; BMI, body mass index; DAS28, disease activity score by 28 joints; Hb, hemoglobin; WBC, white blood cell count; JAKi, Janus kinase inhibitors; y, years.

D. Box plots of plasma (p)Glucose, triglyceride-glucose (TyG) index, and urine (u)C-peptide in RA-SW patients with high (above 157 pmol/L, F, n=81; M, n=39) and low (F, n=122; M, n=27) pInsulin. P-values are obtained by Mann-Whitney test.

### A. DNA content, Gating strategy

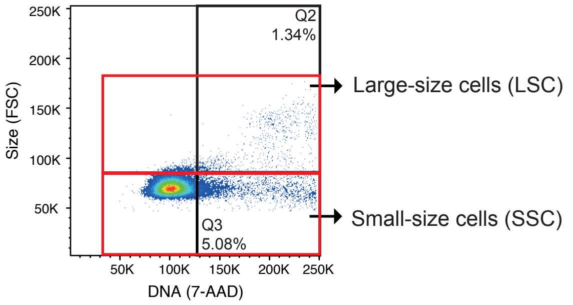


### B. Primers used in qPCR

| Target | Forward (5'to3') | Reverse (5'to3') | Manufacturer |
|---|---|---|---|
| ABL1 | GCTGAGATACGAAGGGAGGG | TGGATAATGGAGCGTGGTGA | Sigma-Aldrich/Merck |
| BIRC5 | GACCACCGCATCTCTACATTC | TGCTTTTTATGTTCCTCTATGGG | Sigma-Aldrich/Merck |
| FGR | GTGCCTACTCCCTGTCCATC | CACAGCCCGTCATTCACCT | Sigma-Aldrich/Merck |
| IFNG | TTTGGGTTCTCTTGGCTGTT | TCCGCTACATCTGAATGACCT | Sigma-Aldrich/Merck |
| INSR | AGATGACAACGAGGAGTGTGG | AGCCGTGTGACTTACAGATGG | Sigma-Aldrich/Merck |
| IRS1 | GTTTCCAGAAGCAGCCAGAG | GGATTTGCTGAGGTCATTTAGG | Sigma-Aldrich/Merck |
| IRS2 | CTTCTTGTCCCACCACTTGA | TGAAACAGTGCTGAGCGTCT | Sigma-Aldrich/Merck |
| PFKFB3 | CCTACAACTTCTTCCGCCCC | CCGCAATTTGTCCCCCTTCT | Sigma-Aldrich/Merck |
| PIK3CG | GGCGACAGACACAATGACAA | GGGTTAGCACAAATGGCACT | Sigma-Aldrich/Merck |
| STAT5A | TGAAGACCCAGACCAAGTTTG | CCATCACGCAGCAGTTGTT | Sigma-Aldrich/Merck |
| SYK | CATCATCAGTCAGAAGCCTCAG | CGTAGGAGCCGTTGTTGTC | Sigma-Aldrich/Merck |
| TBET | CGCCAGGAAGTTTCATTTG | TTATGGAGGGACTGGAGCAC | Sigma-Aldrich/Merck |
| Reference gene | | Assay number | |
| ACTB | | qA-01-0104S | TATAA Biocenter |

C.

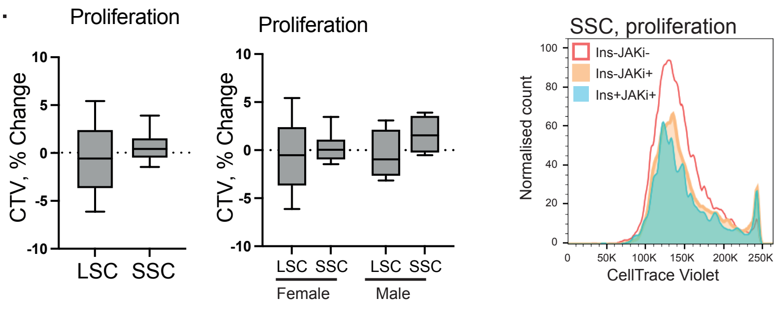


D.

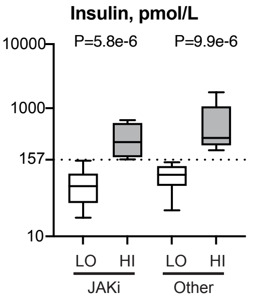


### E. Gene expression in CD4+ cells treated with Insulin

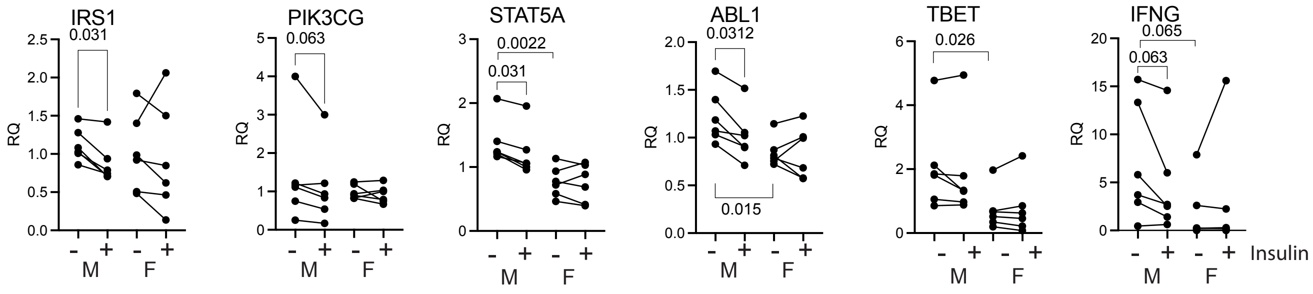


**Supplementary Figure S2.**

A. Gating strategy of CD4+ cells into small-size cells (SSC) and large-size cells (LSC), by forward cell scatter and 7-aminoactinomycin D (7AAD) staining.

B. Sequences of primers used in qPCR.

C. Histogram of CellTrace violette (CTV) dilution in control, insulin and JAKi-treated cells. Box plots of proliferation change by CTV in small size cells (SSC) and large size cells (LSC)

D. Box plots of plasma insulin levels in JAKi-treated (Hi, n=7, Lo, n=17) and other patients (Hi, n=5, Lo, n=27), split by high (Hi) and low (Lo) insulin levels. P-values obtained by Mann-Whitney statistics.

E. Dot plot of gene expression in insulin-treated CD4+ cells, by qPCR. Presented in relative quantity (RQ) to ACTB1. P-values are obtained by paired Wilcoxon test.

A

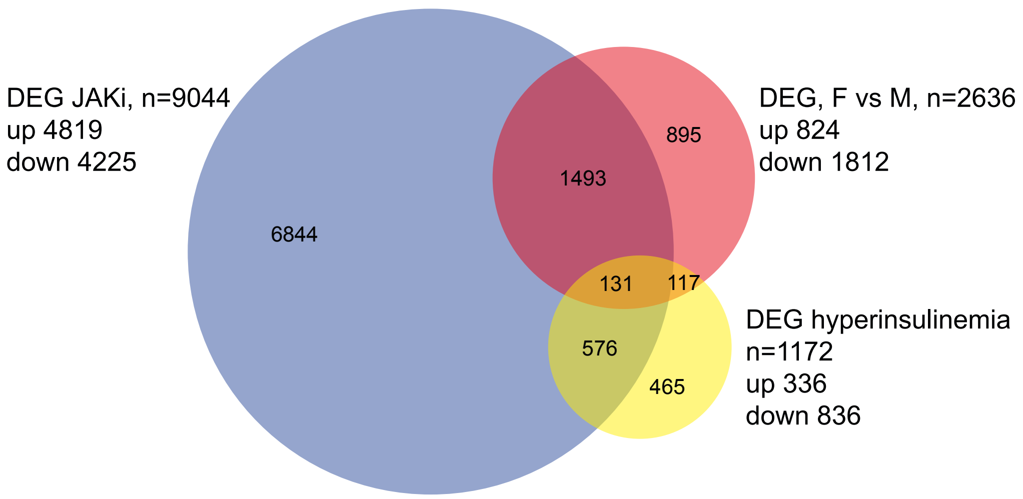


B

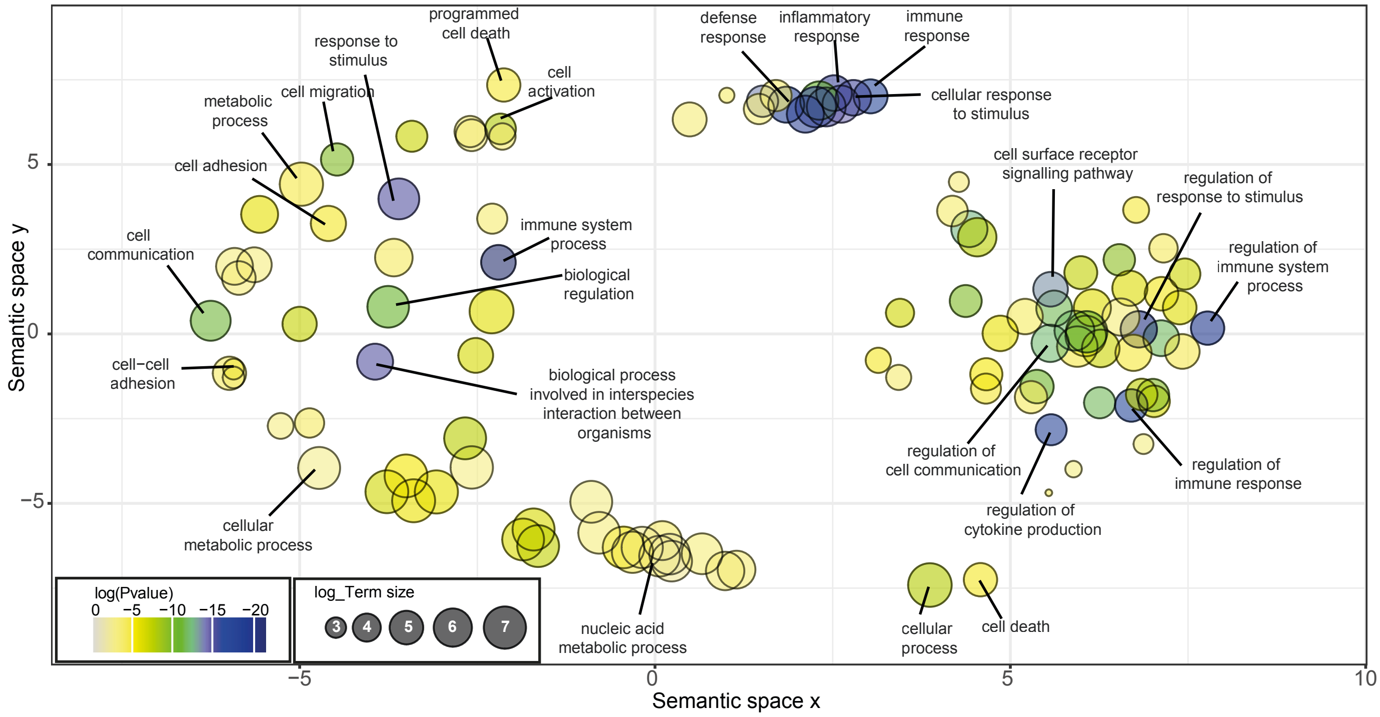


**Supplementary Figure S3.**

A. Venn diagram of differentially expressed genes (DEG, expression basemean >10, nominal p-value<0.05)

B. Semantic plot of the enriched GO:BP (FDR<0.05) identified among common DEG, by Revigo
